# Testing effects of paced breathing on plasma Aβ and brain perivascular spaces

**DOI:** 10.64898/2026.01.08.26343624

**Authors:** Kaoru Nashiro, Jungwon Min, Hyun Joo Yoo, Christine Cho, Martin J. Dahl, Paul Choi, Hye Rynn J. Lee, Jeiran Choupan, Noah Mercer, Padideh Nasseri, Andy Jeesu Kim, Kalekirstos Alemu, Nicole F. Rose, Alexandra Ycaza Herrera, Rachel Custer, Markus Werkle-Bergner, Julian F. Thayer, Lorena Sordo, Elizabeth Head, Mara Mather

## Abstract

Aging is the strongest known risk factor for Alzheimer’s disease (AD), and elevated plasma amyloid-β (Aβ) levels in healthy adults are associated with increased AD risk. Aging is also associated with autonomic imbalance, characterized by increased sympathetic and decreased parasympathetic activity. In our previous randomized clinical trial, we found that four weeks of daily slow-paced breathing designed to enhance parasympathetic activity reduced plasma Aβ42 and Aβ40 levels in younger and older adults and showed a trend toward increasing Aβ42/Aβ40 ratio only in older adults. The primary goal of the current study was to extend these findings in 62 adults aged 50 to 70 years using randomized assignment to 10 weeks of slow-paced breathing or a random-paced breathing control with three assessment time points. Secondary objectives included examining the effects of slow-paced breathing on brain structure (i.e., perivascular space and hippocampal volumes) and cognitive performance. Consistent with prior findings, the slow-paced breathing group showed greater decreases in plasma Aβ42 than the control group.

However, group differences were not significant for Aβ40 or Aβ42/Aβ40 ratios, and no significant effects were observed for the secondary outcomes. The non-significant findings may be due to changes we made to both intervention and control condition methods relative to our previous trial. Further research is needed to explore the underlying mechanisms and potential effects of slow-paced breathing on Aβ accumulation in the brain.

**Highlights:**

- Participants were randomly assigned to slow-placed breathing or a breathing control
- Individualized protocols determined breathing paces
- Ten weeks of daily slow-paced breathing practice reduced plasma Aβ42 levels

## 1. Introduction

Aging is the strongest known risk factor for Alzheimer’s disease (AD), a progressive neurodegenerative disorder. Beginning at age 65, the risk of AD approximately doubles every five years (Ziegler-Graham et al., 2008). Understanding how aging contributes to increased AD risk is essential for developing more effective prevention and treatment strategies (Herrup, 2010). While much previous research has focused on pathological pathways such as oxidative stress (Ionescu-Tucker & Cotman, 2021), cardiovascular risk (Luchsinger & Mayeux, 2004), and inflammation in aging (Pugazhenthi et al., 2017), less attention has been paid to the potential role of changes in autonomic nervous system function.

Aging causes opposing changes in the two major branches of autonomic function (Mather, 2024). Parasympathetic activity, indexed by vagally mediated heart rate variability (HRV), is approximately 80 percent lower at age 60 than at age 20 (Natarajan et al., 2020), whereas sympathetic activity, reflected in part by elevated plasma noradrenaline levels, increases with age, particularly during sleep. For example, older men exhibit roughly 75 percent higher noradrenaline concentrations during sleep compared to younger men (Prinz et al., 1979).

Our hypothesis (Mather, 2025) is that these autonomic changes during young and middle adulthood contribute to age-related increases in Aβ42 levels (Luo et al., 2022; Toledo et al., 2013; Zecca et al., 2021), which in turn are associated with greater AD risk (Jensen et al., 1999; Song et al., 2011; Zetterberg, 2022). Aβ is generated when amyloid precursor protein (APP) is sequentially cleaved by β and γ secretases (Nunan & Small, 2000). Animal studies suggest that increased noradrenergic activity promotes Aβ production: activating α2a adrenergic receptors increases Aβ production by promoting β-secretase cleavage of APP (Chen et al., 2014) while activating β2 adrenergic receptors increases Aβ production by enhancing γ secretase activity (Ni et al., 2006). Studies with rodent models reveal that propranolol and carvedilol, β-blockers that cross the blood-brain barrier (BBB), reduce cognitive impairments and reduce brain Aβ levels (Dobarro, Gerenu, et al., 2013; Dobarro, Orejana, et al., 2013a, 2013b; Wang et al., 2011).

Consistent with these rodent studies, in a study of 69,081 Danish patients treated for hypertension, those treated with propranolol and carvedilol had lower risk of Alzheimer’s disease than those treated with β-blockers that had low BBB permeability (Beaman et al., 2023).

In contrast with the β- and γ-secretase pathway, if APP is cleaved instead by α secretase, Aβ formation is prevented. Cholinergic muscarinic receptors activate the α-secretase APP nonamyloidogenic pathway and suppress the β-secretase amyloidogenic pathway (Cho et al., 2022; Giacobini et al., 2022; Hock et al., 2000; Nitsch et al., 2000). This is relevant for age-related autonomic changes as parasympathetic vagus nerve activity stimulates muscarinic receptors in the brain (Nichols et al., 2011). Thus, age-related increases in sympathetic activity and decreases in parasympathetic activity could both bias towards increased Aβ production, via increased noradrenergic and decreased muscarinic receptor activity, respectively. Autonomic shifts with aging may also impair Aβ clearance. In the brain, noradrenaline reduces the effectiveness of the glymphatic waste clearance system by shrinking the interstitial space, thereby limiting the removal of protein waste products such as Aβ (Hussain et al., 2023; Xie et al., 2013). Glymphatic clearance is most active during sleep (Mendelsohn & Larrick, 2013; Xie et al., 2013), when noradrenaline levels are low. Therefore, sustained elevations in tonic noradrenaline with aging may reduce clearance capacity and promote Aβ accumulation. Interventions that reduce noradrenaline levels may help support glymphatic function by decreasing neuronal cell body volume, expanding interstitial space, and facilitating fluid flow.

One possible strategy to reduce tonic noradrenergic activity is to stimulate the baroreflex, a homeostatic feedback mechanism that regulates arterial blood pressure. When baroreceptors sense elevated pressure, the reflex enhances parasympathetic (vagal) activity and inhibits sympathetic tone, including output from the locus coeruleus (Murase et al., 1994; Schneider et al., 1995), the brain’s primary source of noradrenaline. Baroreflex activity can be enhanced through slow-paced breathing at a frequency of approximately 0.1 Hz (Bernardi et al., 2001; Radaelli et al., 2004). Slow-paced breathing also increases heart rate oscillations through respiratory HRV (i.e., the acceleration of heart rate during inhalation and deceleration during exhalation; Menuet et al., 2025).

In our previous clinical trial (Nashiro et al., 2023; Yoo et al., 2023), both younger and older participants assigned to daily slow-paced breathing to increase heart rate oscillations (Osc+) showed reduced plasma Aβ42 and Aβ40 levels after four weeks (Min et al., 2023). In contrast, those in a condition that attempted to minimize oscillations (Osc-) showed increases in both Aβ42 and Aβ40 levels. Critically, plasma Aβ42/Aβ40 ratio showed a trend toward an interaction effect only in older adults, for whom the ratio increased for Osc+ relative to Osc−. Prior research suggests that lower Aβ42/40 ratios are associated with higher brain amyloid pathology because as older adults’ brain clearance mechanisms decline and their brain Aβ aggregation increases, Aβ42 is less likely than Aβ40 to be cleared to the brain to the periphery. Our findings suggested that the Osc+ intervention promoted clearance from the brain to the periphery of Aβ42 in older adults in whom brain waste clearance processes are starting to decline, but not in younger adults in whom brain waste clearance is still effective. In a separate study with younger adults (Nashiro et al., 2025), one week of daily slow-paced mindfulness breathing reduced plasma Aβ, whereas mindfulness with normal breathing led to increases in plasma Aβ. No changes were observed in a third no-intervention control group (Nashiro et al., 2025). As expected given the young age of the cohort (ages 18-35), none of the three groups showed changes in plasma Aβ42/Aβ40 ratio.

In the current clinical trial (Nashiro et al., 2024), we aimed to extend these findings in adults aged 50 to 70 years and expand the study by implementing a longer, 10-week intervention with three assessment time points and by more rigorously identifying individually optimal breathing paces. As in our prior studies, plasma Aβ42 and Aβ40 levels, along with the Aβ42/40 ratio, served as the primary outcomes. To explore underlying mechanisms, we used MRI to examine perivascular space (PVS) volume, a secondary outcome. PVS dilation is a proposed marker of impaired glymphatic clearance and is associated with increased dementia risk in aging (Bouvy et al., 2020; Kim et al., 2020; Mestre et al., 2017). We also assessed hippocampal volume and cognitive performance (measured by Lumosity brain games that are designed to improve various core cognitive skills including memory, attention, mental flexibility and reasoning; Supplementary Table 1) as additional secondary outcomes to evaluate broader neurobiological and behavioral effects of the intervention. This Heart rate and Breathing Effects on Attention and Memory (‘HeartBEAM’) study was registered on ClinicalTrials.gov (NCT05602220).

## 2. Methods

### 2.1. Participants

We enrolled 91 healthy adults between the ages of 50 and 70 (Fig. 1). Recruitment efforts included the use of flyers, emails, online posting websites, University of Southern California (USC) Healthy Minds research volunteer registry, and a third-party recruitment firm called TrialFacts who was contracted to provide referrals for the study. Prospective participants who were fluent in English, non-pregnant and non-menstruating for at least the past year, who had normal or corrected-to-normal vision and hearing, who had a home computer with a physical keyboard, who had access to reliable internet, who agreed to provide blood and urine samples, who weighed at least 110 pounds, who agreed to devote up to 60 minutes daily for 12 weeks, who had an email account that was regularly checked, and who had a phone that receives text messages were screened and invited to participate. We excluded those who had a condition that would impede completing the HRV biofeedback intervention (e.g., abnormal cardiac rhythm, heart disease including coronary artery disease, angina and arrhythmia, self-reported cognitive impairment, dyspnea), who regularly practiced any relaxation, meditation or yoga that involved breathing-focused practices lasting for more than an hour each week, who had regularly played Lumosity games in the past 6 months, who had any conditions that are not safe for MRI (e.g., metals in the body, claustrophobia, cardiac pacemaker), and who had previously participated in heart rate variability biofeedback studies in the USC Emotion and Cognition Lab. Upon passing the screening, participants completed the consent form approved by the USC Institutional Review Board and created an account on the study application. Participants received compensation for completing home assessments, lab visits and the intervention (for details, see ‘Participant Compensation in the Supplemental Materials). Twenty-nine participants withdrew from the study prior to randomization (Fig. 1; Tables 1 and 2).

**Fig. 1.**
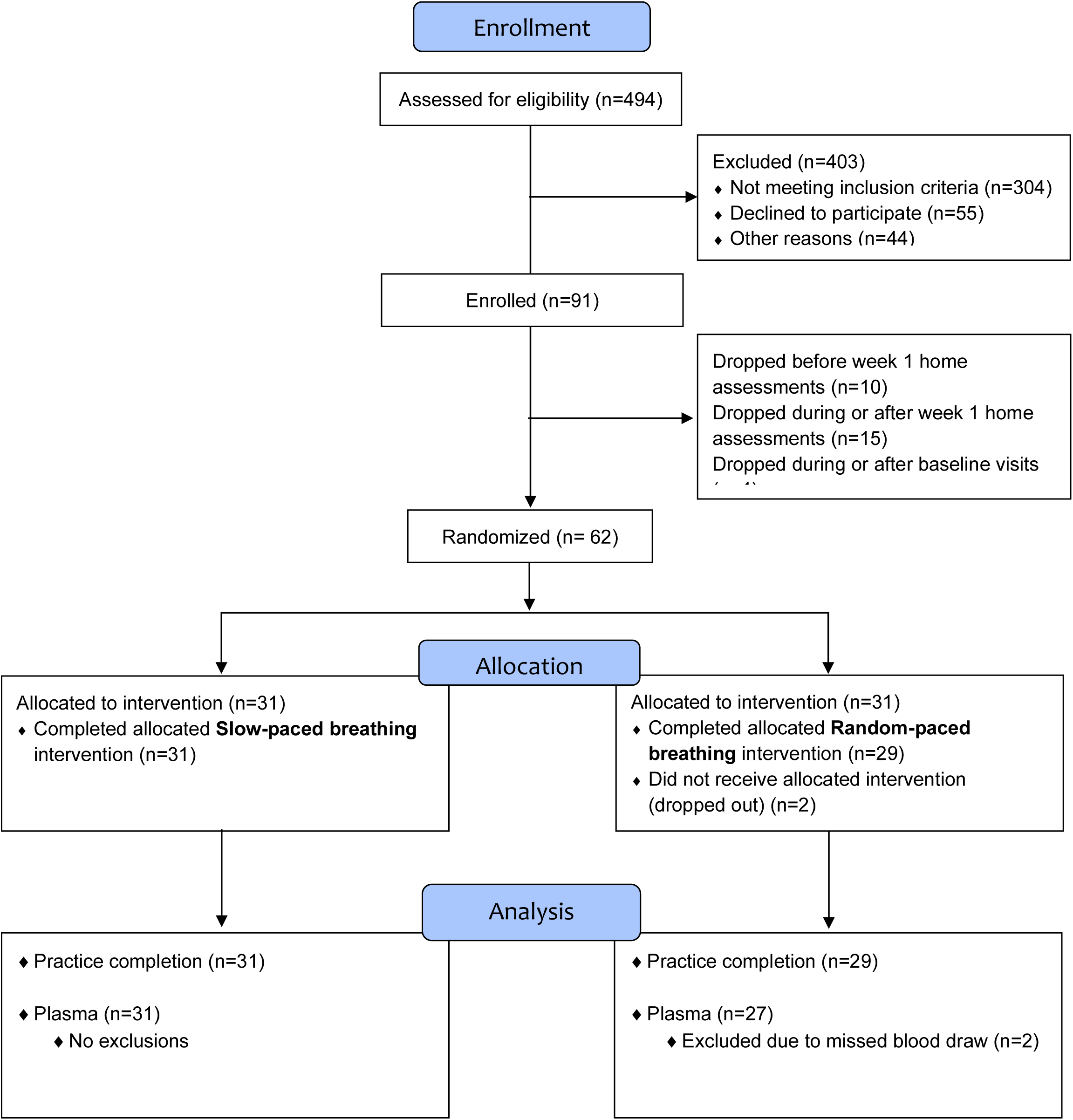
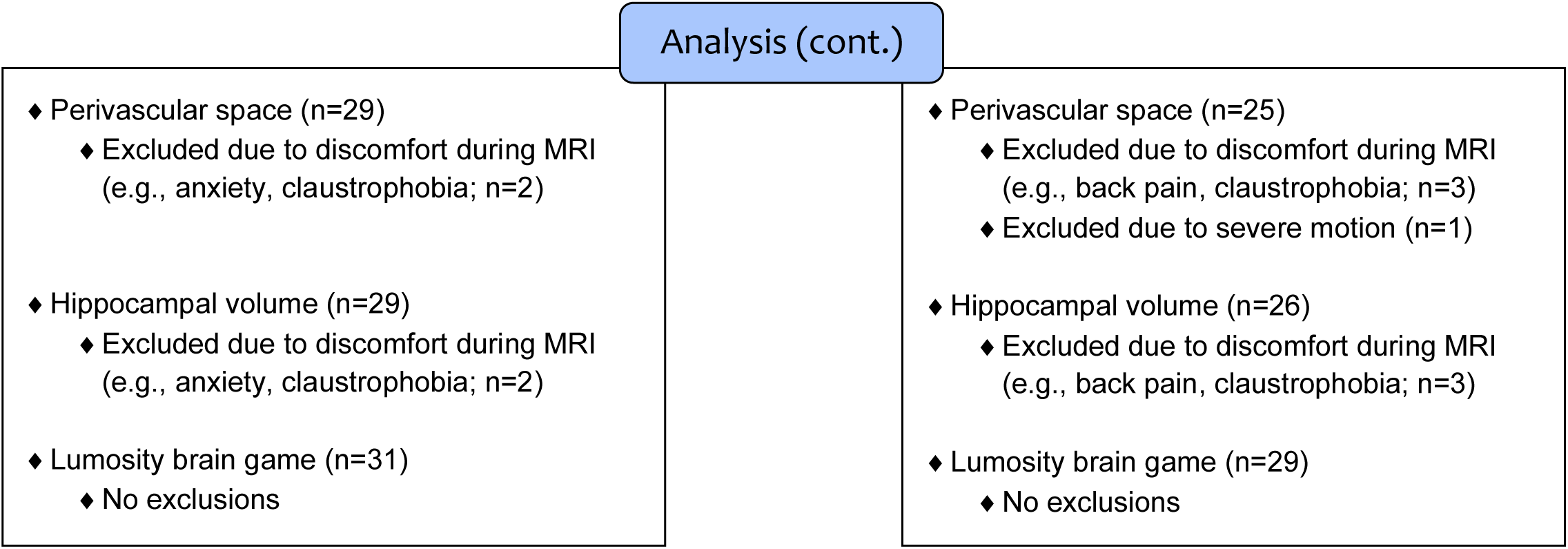
Flow diagram. Numbers of participants in each intervention condition, how many participants completed each measure, and how many were included vs. excluded in each analysis.

**Table 1.**
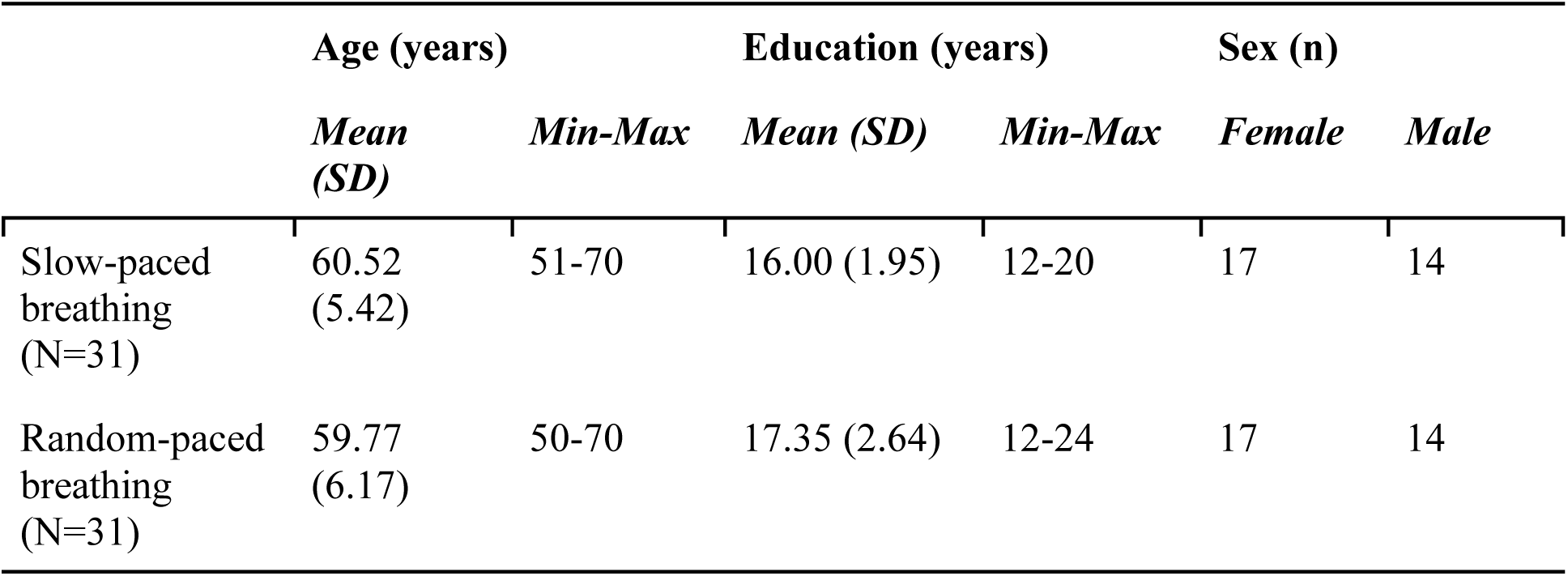
Participants’ mean age and education, and number of participants by sex.

|  | Age (years) |  | Education (years) |  | Sex (n) |  |
| --- | --- | --- | --- | --- | --- | --- |
|  | *Mean (SD)* | *Min-Max* | *Mean (SD)* | *Min-Max* | *Female* | *Male* |
| Slow-paced breathing (N=31) | 60.52 (5.42) | 51-70 | 16.00 (1.95) | 12-20 | 17 | 14 |
| Random-paced breathing (N=31) | 59.77 (6.17) | 50-70 | 17.35 (2.64) | 12-24 | 17 | 14 |

**Table 2.** Participant race and ethnicity.

|  | Slow-paced breathing (N=31) | Random-paced breathing (N=31) |
| --- | --- | --- |
| **Hispanic** |  |  |
| African-American | 1 | 1 |
| Biracial | 0 | 1 |
| Caucasian | 3 | 2 |
| Prefer not to state | 0 | 3 |
| **Non-Hispanic** |  |  |
| African-American | 1 | 4 |
| Asian | 7 | 5 |
| Biracial | 3 | 0 |
| Caucasian | 15 | 14 |
| **Prefer not to state** |  |  |
| Biracial | 0 | 1 |
| Prefer not to state | 1 | 0 |

### 2.2. Overview of 12-week protocol schedule

Over the course of the study, participants completed two weeks of home assessments, five lab visits, and a 10-week home intervention. See Fig. 2 for an outline of the weekly study schedule.

**Fig. 2.**
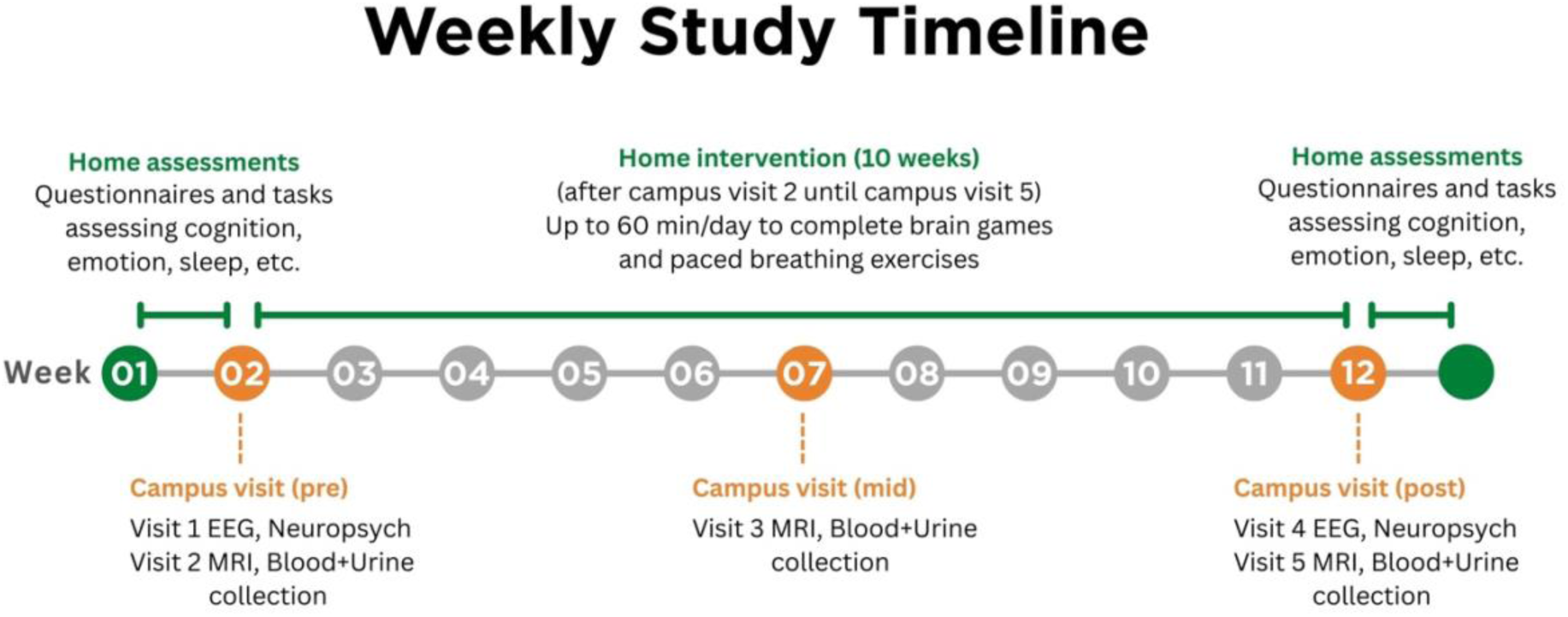
Overview of weekly study schedule over 12 weeks.

#### 2.2.1. Home assessments

Week 1 of the study was completed from home, which consisted of 45 minutes of baseline cognitive and emotion assessments daily over six consecutive days (Nashiro et al., 2024 for a complete list of assessments). These assessments were repeated during Week 12, again from home, after the participant had completed all lab visits and the 10-week intervention.

#### 2.2.2. Lab visits

Participants attended a total of five lab visits: two visits in Week 2 (pre), one visit in Week 7 (mid), and two visits in Week 12 (post). For Visit 1, participants completed questionnaires assessing sleep and functional activities as well as neuropsychological assessments, which consisted of Trail Making Test A/B, Animal Fluency, Multilingual Naming Test (MiNT), and the Montreal Cognitive Test (MoCA). Participants then completed a baseline EEG session (note: EEG outcomes will be reported elsewhere). For Visit 2, participants provided urine and blood samples, completed mood, stress, and sleep questionnaires, and underwent a baseline brain MRI scan. At the end of this visit, participants received their study laptop and ear sensor, which were used to complete two baseline pulse measurements during a 5-minute resting period and a 4-s paced breathing exercise. Participants followed a pacer, breathing in and out through their nose at 15 breaths per minute. Visit 3 occurred at the midpoint of the study at Week 7, and the same tasks from Visit 2 were repeated excluding the resting pulse measurement and paced breathing exercise. For Visit 4, we administered the same tasks from Visit 1 with the exception of the functional activities questionnaire, MiNT and MoCA. Similarly, for Visit 5, the same tasks were repeated as in Visit 2, and the study laptops and ear sensors were collected from participants. Please note that this paper focuses on reporting pre-registered primary and secondary outcome measures (https://clinicaltrials.gov/study/NCT05602220).

#### 2.2.3. Home intervention

Participants completed 10 weeks of intervention at home using their lab-issued study laptop and ear sensor. For the first week of the intervention (Week 2 of the study), participants played a set of Lumosity brain games (Supplementary Table 1) and completed a 5-minute resting pulse measurement daily. From Week 3 to Week 12, participants played a set of Lumosity brain games and completed two sessions of 15-minute paced breathing practices. The Lumosity brain games examined different cognitive domains including memory, attention, mental flexibility, reasoning, processing speed, spatial orientation, and problem skills (Steyvers & Schafer, 2020). Participants played two sets, and each set consisted of six games. The sets of brain games alternated every day.

Participants were randomly assigned to one of two conditions: slow-paced breathing condition and random-paced breathing condition (for more details, see ‘Randomization’ in the Supplementary Materials). Heart rate was monitored during breathing practices with a sensor placed on the earlobe, which connected to a USB device that plugged into the study laptop. The laptop screen displayed real-time HRV biofeedback with a visual pacer that showed a ball moving up, down, and horizontally along a line. Participants inhaled when the ball went up, exhaled when it went down, and paused their breath when it moved horizontally. An auditory sound was also provided as a guide. Before each breathing practice, participants were advised to sit upright in a chair with their feet flat on the floor and hands resting on their lap. Each day, they completed two sessions of 15-minute breathing practices.

##### 2.2.3.1. Slow-paced breathing condition

Participants in this slow-paced breathing condition were guided through breath paces at 10, 12, and 15 seconds per breath. This protocol was adapted from our previous HRV-ER trial (Min et al., 2023), which used a standard HRV-biofeedback method where participants tested various breathing rates and selected one based on various features (such as spectral frequency power around the breathing frequency) posited to be associated with resonance produced by breathing at the same frequency as the person’s inherent baroreflex frequency (Shaffer et al., 2014). Results showed that both younger and older adults breathing at 11, 12, or 13 seconds per breath had greater reductions in plasma Aβ compared to those at 9 or 10 seconds. To explore whether slower rates were more effective, in this study we included a 15-second pace. We also improved the rigor of the pace selection method. Our previous HRV-ER study relied on a single test trial each week and could be influenced by order effects or variability. In the current study, we had participants practice multiple paces repeatedly at home until we could statistically discriminate the effectiveness of the paces in generating large heart rate oscillations. Poorly performing paces were removed from the practice set until there was just one best pace left, at which point we added two additional paces, one a little faster and the other a little slower than the best pace (Nashiro et al., 2024). During daily sessions, participants received a “relaxation score.” A greater score indicated higher amplitude of heart rate oscillatory activity ranging in the 0.04 to 0.26 Hz frequency (equivalent to 4–25 seconds/breath). Participants were encouraged to breathe diaphragmatically to maximize their score.

##### 2.2.3.2. Random-paced breathing condition

Participants in this condition were guided through breathing paces at 4, 5, and 6 seconds per breath, which approximate typical breathing rates with somewhat randomized inhalation and exhalation lengths to avoid generating rhythmic heart rate oscillations. While completing these exercises, participants received an “alertness score.” A high score indicated a decrease of heart rate oscillatory activity ranging in the 0.04 to 0.26 Hz frequency. Participants were encouraged to earn high alertness scores by breathing naturally and lightly, but not too deeply.

#### 2.2.4. MRI scan parameters

We used a 3T Siemens MAGNETOM Trio scanner with a 32-channel head array coil at the USC Dana and David Dornsife Neuroimaging Center. All scans were acquired pre-, mid- and post-intervention. A high-resolution, T1-weighted magnetization prepared rapid acquisition gradient echo (MPRAGE) scan was acquired with TR = 2,400 ms, TE = 2.34 ms, slice thickness = 0.70 mm, flip angle = 8°, field of view = 224 mm, and voxel size = 0.7 × 0.7 × 0.7 mm (acquisition time 7:40 min). A high-resolution, T2-weighted image was obtained with TR = 3,200 ms, TE = 566 ms, slice thickness = 0.70 mm, field of view = 224 mm, and voxel size = 0.7 × 0.7 × 0.7 mm (acquisition time 8:27 min). Fluid-Attenuated Inversion Recovery (FLAIR) images were collected with TR = 4,800 ms, TE = 441 ms, slice thickness = 1.20 mm, field of view = 256 mm, and voxel size = 1.0 × 1.0 × 1.2 mm (acquisition time 5:28 min). T2-weighted high-resolution hippocampal imaging was acquired with TR = 6,490 ms, TE = 16 ms, slice thickness = 2.0 mm, flip angle = 120°, field of view = 206 mm, and voxel size = 0.4 × 0.4 × 2.0 mm (acquisition time 4:15 min). In addition, we acquired proton density-weighted whole-brain (low-res) and partial field of view (high-res) sequences with magnetization transfer preparation pulse for locus coeruleus visualization (results from the locus coeruleus scans will be reported elsewhere).

### 2.3. Analyses

#### 2.3.1. Heart rate variability

During the first week of the intervention (Week 2 in Figure 2), participants completed daily 5-minute resting HRV recordings, including an initial seated baseline assessment during their first lab visit. During this initial baseline lab visit only, a fixed 4-s paced breathing condition was included to control resting breathing and to enable estimation of participants’ individual baroreflex frequency (Sakakibara et al., 2020), or frequency at which the cardiac baroreflex naturally oscillates (P. M. Lehrer et al., 2003). Beginning in the second week of the intervention (Week 3 in Figure 2), when HRV biofeedback training began, participants performed two 15-minute breathing sessions per day. In all sessions (whether resting or training) participants wore an infrared pulse plethysmograph (PPG) ear sensor connected to the emWave Pro system (HeartMath® Institute, 2020), which recorded pulse wave data at a sampling rate of 370 Hz. Each 15-minute training session was divided into three 5-minute segments, each involving a different paced breathing rate. Participants followed on-screen breathing pacers while receiving real-time heart rate feedback. Although the feedback was based on the same coherence score calculation (described below), it was displayed as a *relaxation score* in the slow-paced breathing condition and as an *alertness score* in the random-paced breathing condition.

To estimate each participant’s individual baroreflex frequency, spectral peak detection was performed specifically on the inter-beat interval data obtained during the fixed 4-s paced breathing condition at the initial baseline lab visit <u>(e.g., Sakakibara et al. 2020)</u>. Power spectra were computed using the Lomb–Scargle periodogram and examined within the putative baroreflex frequency range (0.067–0.15 Hz). The first peak within this range was extracted for each 5-minute baseline segment and used as an individualized index of spontaneous low-frequency heart-rate oscillatory activity.

For each 5-minute segment of both the resting and training sessions, a coherence score was calculated using the emWave software. This score was defined as peak power divided by (total power – peak power), with peak power determined as the highest spectral peak within the 0.04–0.26 Hz range, computed by integrating a ±0.015 Hz window. Total power was computed over the 0.0033–0.4 Hz range. In the slow-paced breathing condition, the coherence score was presented as a relaxation score; in the random paced breathing condition, it was reverse-coded and displayed as an alertness score (i.e., alertness score = [coherence × −1] + 10).

Further HRV analysis was conducted using the RHRV package in R (Martínez et al., 2017). Inter-beat interval (IBI) data extracted from the emWave system were preprocessed to remove ectopic beats and artifacts. HR was computed and interpolated at 4 Hz, and the signal was filtered using standard RHRV settings. Time-domain indices, such as mean heart rate and root mean square of successive differences (RMSSD), were computed from the cleaned data. In addition, frequency-domain indices were derived using Fast Fourier Transform (FFT), and spectral power was calculated within standard HRV frequency bands: low frequency (LF: 0.04–0.15 Hz) and high frequency (HF: 0.15–0.4 Hz). These HRV indices and coherence scores were then used in subsequent statistical analyses. Before conducting the analyses, all HRV-related variables were tested for normality using the Shapiro–Wilk test. The results indicated that all variables except for mean heart rate significantly deviated from a normal distribution.

Specifically, RMSSD, LF power, HF power, and coherence scores failed to meet the assumption of normality and were therefore transformed using the natural logarithm. However, even after log transformation, several variables—including coherence scores at rest and training, RMSSD, and HF power—continued to violate normality assumptions (*p* < .05). To account for this, we employed linear mixed-effects models, which are more robust to violations of normality assumptions, to assess the effects of time (rest, training), condition (slow-paced, random-paced), and their interaction. Analyses were conducted in R (version 4.3.1) using the lme4 and lmerTest packages. The model included fixed effects of time, condition, and their interaction, as well as a random intercept for each participant to account for repeated measures.

Fixed effects were tested using Type III ANOVA, with partial eta squared (*η²*) reported as the measure of effect size. Post-hoc pairwise comparisons were performed using the emmeans package with Tukey adjustment, and Cohen’s *d* was calculated for each contrast.

### 2.3.1. Blood assays

Blood draws were conducted at the same time of the day across the three visits for each participant, and participants were not asked to fast. A phlebotomist attempted to draw 16.5 ml of blood from each participant’s arm into three tubes using antecubital venipuncture. When the arm was not viable, venipuncture was performed on the hand or wrist instead. To separate plasma from red blood cells, the whole blood from the K2 EDTA tubes was centrifuged at the speed of 1500 RPM for 15 min at room temperature (15°C). Plasma was aliquoted in cryovials and stored at −80 °C. The frozen plasma samples were transferred and assayed at University of California, Irvine. Quanterix assays were used to measure plasma Aβ40 and 42 (Simoa Neurology 3-Plex A Advantage Kit).

#### 2.3.2. PVS volume

PVS was automatically segmented and quantified using processing methods described previously (Sepehrband et al., 2019). Using HCP pipelines (Glasser et al., 2013; Milchenko & Marcus, 2013; Sotiropoulos et al., 2013), T1w and T2w images were corrected for gradient nonlinearity, readout, and bias field, aligned to AC-PC subject space, and registered to MNI 152 space using FSL’s FNIRT (Glasser et al., 2013). Individual white and pial surfaces were then generated using the FreeSurfer software and the HCP pipelines. Preprocessed T1w and T2w images were filtered using adaptive non-local mean filtering technique. To increase visibility of the PVS, an enhanced PVS contrast (EPC) was obtained by dividing the T1w image by the T2w image (Sepehrband et al., 2019). A frangi filter was applied to the EPC to identify vesselness probability for each voxel to guide the segmentation of the tubular PVS structures. The vesselness probability map was applied with thresholds of p < 4e-6 in the white matter and the basal ganglia, optimized for this dataset based on multiple experts fine-tuning. The thresholded map was then binarized to obtain PVS voxels. PVS voxels are sometimes hard to distinguish from white matter hyperintensities (WMH) that mimic PVS morphology, leading to false positives (Barisano, Lynch, et al., 2022; Sepehrband et al., 2021). Thus, we used FLAIR images (or T1w images when FLAIR was not available) to create WMH masks and remove PVS voxels falling within the WMH masks. WMH masks were generated by the Sequence Adaptive Multimodal Segmentation tool (Cerri et al., 2021) and the Lesion Growth Algorithm (Schmidt et al., 2012) as this was shown to improve sensitivity and specificity of PVS segmentation (Barisano, Sepehrband, et al., 2022; Sepehrband et al., 2021). In addition, the PVS volumes were manually inspected by two trained researchers, and false positives were removed. PVS volumes were extracted from the centrum semiovale (CSO), which was a main region of interest (ROI) since it has been historically used as the clinical ROI to assess PVS. CSO PVS volume was calculated by adding up PVS volumes of each of the regions known to comprise the CSO. The CSO mask was generated based on the ROIs from Desikan-Killiany atlas listed in Supplementary Table 2. CSO volume fraction was then obtained by dividing the total PVS volumes of these added regions by the corresponding white matter volumes. PVS volumes were extracted from other brain regions based on the Freesurfer’s Desikan-Killiany-Tourville adult cortical parcellation atlas (Klein & Tourville, 2012). PVS volume fractions were then calculated by dividing the volume of the PVS in the region by the white matter volume of that same region.

#### 2.3.3. Hippocampal volume

The preprocessed T1w images used for the PVS analysis were also utilized in the hippocampal volume analysis. FreeSurfer (v7.3.1) was employed for cortical parcellation and subcortical segmentation of the T1w volume, including total intracranial volume (TIV) estimation for head size correction (surfer.nmr.mgh.harvard.edu; Dale et al., 1999; Fischl et al., 2002). The FreeSurfer processing workflow included non-brain tissue removal, automated Talairach transformation, intensity normalization, segmentation, tessellation of the gray/white matter boundary, topology correction, and surface deformation. Additionally, T2w images were integrated into the FreeSurfer pipeline to refine the pial surface by distinguishing dura and vasculature.

After the initial FreeSurfer processing, FreeSurfer’s longitudinal processing pipeline was applied to enhance within-subject consistency across time points (Reuter et al., 2012). This approach involves creating an unbiased within-subject template using the available timepoints, which serves as a reference to improve the accuracy and reliability of longitudinal measurements. The template is then used to initialize each timepoint, reducing segmentation variability and increasing statistical power in detecting cortical and subcortical changes over time.

Following the completion of the FreeSurfer processing pipeline, which incorporated both T1w and T2w images, hippocampal subfield segmentation was performed using FreeSurfer’s hippocampal subfield segmentation tool (v22) in version 7.3.1 (Iglesias et al., 2015). This segmentation process utilized the outputs from the FreeSurfer pipeline and was further refined by incorporating a dedicated high-resolution hippocampal scan to enhance anatomical accuracy. The method leverages probabilistic maps derived from high-resolution ex vivo MRI to initially segment each hippocampus into 19 subregions, which are subsequently merged based on predefined anatomical schemes. One participant was unable to complete the high-resolution hippocampal scan at the mid time point; therefore, for that participant segmentation based solely on T1w and T2w images was used at all three time points to maintain consistency.

We applied quality control procedures based on the guidelines for FreeSurfer-based segmentation of hippocampal subfields (Sämann et al., 2022). First, we identified outliers exceeding ±2 standard deviations (approximately 5% of the sample distribution) for each subfield volume, as well as for total brain volume, total gray matter (GM) volume, intracranial volume (ICV), and the GM/ICV ratio. Second, we calculated deviations in the rank order of subfield volumes within each participant, comparing them to normative patterns derived from several large-scale datasets (Sämann et al., 2022). Additionally, histograms of each measure were visually inspected. Finally, all datasets were reviewed using the FreeSurfer-generated HTML reports in a standard browser, with particular attention paid to the datasets flagged in previous steps. Visual inspection followed established quality control criteria: (1) Is the binary hippocampal mask clearly visible? (2) Are large portions of the hippocampus missing or truncated? (3) Are there any abnormalities specific to individual subfields? No datasets were excluded as a result of this QC process, and no manual edits were performed to avoid introducing potential bias.

#### 2.3.4. Lumosity brain training performance

Participants played 12 brain training games through the Lumosity platform (https://www.lumosity.com/) from 6 cognitive domains, Attention, Flexibility, Language, Math, Memory, Reasoning (see Supplementary Table 1 and Steyvers & Schafer, 2020). Games were assigned to one of two daily alternating sets. On average, participants played each game 32 to 34 times over the course of 10 weeks (Supplementary Table 3).

Our analyses focused on raw performance scores that were standardized within each game across participants and game plays before analyses. Over the course of the study, performance on each of the games gradually improved, following a smooth learning curve (Supplementary Figure 1). Average performance was positively coupled over games, indicating a shared latent construct (Supplementary Figure 2). Thus, intervention effects were evaluated on a latent level, aggregating over individual games.

To this end, game performance time series were down-sampled within each game by averaging over three adjacent game plays (bins: 1- 4, 4-7, 7-10, […] 28-31) to counteract missingness of individual time points (Supplementary Figure 3) and to reduce noise. Remaining missing values were interpolated using adjacent time points (on a participant level; using *inpaint_nans* in Matlab).

Next, Task Partial Least Squares Correlation (T-PLSC) was employed to estimate latent performance trajectory of cognitive change over the intervention (Krishnan et al., 2011). T-PLSC is a multivariate statistical technique used to identify latent variables that optimally capture the shared variance between two data sets—in this case, task performance in the 12 games and an experimental design matrix (i.e., the 10 time bins) reflecting the duration of the intervention.

Unlike univariate approaches, T-PLSC analyzes the covariance structure across all games simultaneously, enabling detection of patterns that reflect coordinated changes in performance. This method is especially suited for high-dimensional data, as it reduces the dimensionality while preserving game interdependence. In the present study, T-PLSC was applied to determine whether changes in performance across multiple Lumosity games were modulated by the intervention, capturing latent effects that might be obscured when games are analyzed in isolation. The statistical significance of the latent variable was assessed using permutation testing (npermutations = 10000). Specifically, the order of time bins was randomly permuted to generate a null distribution of singular values. The *p*-value was computed as the proportion of permuted singular values that exceeded the original singular value, providing a non-parametric estimate of the likelihood that the observed multivariate association occurred by chance. To assess the stability and interpretability of individual game contributions (saliences or loadings) to the latent variable, bootstrap resampling was conducted (nbootstraps = 10000). In each iteration, participants were resampled with replacement, and the task saliences were re-estimated. The resulting distribution of bootstrap saliences was used to calculate standard errors, from which bootstrap ratios (salience divided by its standard error) were derived. Tasks with higher bootstrap ratios (|BSR|>3) were considered reliable contributors to the latent pattern. Finally, latent performance trajectories were compared across intervention groups by means of time bin-wise independent samples *t*-tests.

## 3. Results

### 3.1. HRV during home training

A linear mixed-effects model was conducted on the log-transformed coherence scores, with time (resting, training) as a within-subjects factor and condition (slow-paced breathing, random-paced breathing) as a between-subjects factor (N = 61; slow-paced: n = 31, random-paced: n = 30). The analysis revealed a significant main effect of time, *F*(1, 59) = 61.05, *p* < .001, *ηp²* = .509, and a significant main effect of condition, *F*(1, 59) = 67.46, *p* < .001, *ηp²* = .533. There was also a significant time × condition interaction, *F*(1, 59) = 79.99, *p* < .001, *ηp²* = .576. Post hoc Tukey-adjusted comparisons showed that coherence significantly increased from rest to training in the slow-paced breathing condition, *t*(59) = –11.95, *p* < .001, *d* = 3.03, 95% CI [–3.67, –2.39]. In the random-paced breathing condition, the change from rest to training was not significant, *t*(59) = 0.79, *p* = .857, *d* = 0.20, 95% CI [–0.31, 0.72]. At rest, there was no significant difference between slow-paced breathing and random-paced breathing conditions, *t*(102.66) = 1.89, *p* = .241, *d* = 0.62, 95% CI [–0.04, 1.27]. However, during training, coherence was significantly higher in the slow-paced breathing condition compared to the random-paced breathing condition, *t*(102.66) = 11.79, *p* < .001, *d* = 3.86, 95% CI [3.04, 4.68]. These increases in coherence during training indicate that participants in the slow-paced breathing condition successfully engaged in the intended protocol and increased their heart rate oscillatory activity.

Building on these findings, we also conducted a linear mixed-effects model on the log-transformed LF power to assess whether similar effects were observed in this standardized frequency domain (0.04 ∼ 0.15 Hz) that covers most of slow breathing frequencies (four participants had at least one segment with a breathing rate below 0.04 Hz or 2.4 breaths per min but none had a pace below 0.04 Hz for their weekly best pace). The model included state (resting, training) as a within-subjects factor and condition (slow-paced breathing, random-paced breathing) as a between-subjects factor. The analysis revealed a significant main effect of state, *F*(1, 59) = 94.10, *p* < .001, *ηp²* = .615, and a significant state × condition interaction, *F*(1, 59) = 7.68, *p* = .007, *ηp²* = .115. Post hoc Tukey-adjusted comparisons showed that LF power significantly increased from rest to training in the slow-paced breathing condition, *t*(59) = –8.89, *p* < .001, *d* = 2.26, 95% CI [–2.84, –1.68], while a smaller but significant increase was also observed in the random-paced breathing condition, *t*(59) = –4.86, *p* < .001, *d* = 1.25, 95% CI [–1.79, –0.72]. Between-group comparisons revealed no significant difference during rest, *t*(99.17) = 0.11, *p* = .999, *d* = 0.04, 95% CI [–0.64, 0.71], but LF power during training was significantly higher in the slow-paced breathing condition compared to the random-paced breathing condition, *t*(99.17) = 3.05, *p* = .015, *d* = 1.04, 95% CI [0.35, 1.73]. These findings further support that participants in the slow-paced breathing condition engaged effectively in the intended training, as reflected by increases in both coherence and LF power.

In contrast, no significant state × condition interactions were observed for heart rate, log RMSSD, or log HF power, indicating that these indices did not show differential change relative to rest in the slow-paced breathing condition versus the random-paced breathing condition during their home practice sessions. The estimated marginal means and standard errors for these HRV indices are reported in Supplementary Table 4.Across participants, individually estimated baroreflex frequencies were generally distributed in the vicinity of 0.1 Hz. During the initial baseline assessment, the first spectral peak within the baroreflex frequency range (0.067–0.15 Hz) extracted from the 4-s paced breathing session was centered near 0.1 Hz across both breathing conditions. Specifically, the mean peak frequency was 0.094 Hz in the slow-paced breathing condition and 0.096 Hz in the random-paced breathing condition, yielding an overall mean of 0.095 Hz. These results indicate consistent clustering of estimated individual baroreflex frequencies around ∼0.1 Hz across participants.

### 3.2. Blood assays

For both plasma Aβ42 and Aβ40, we observed an unexpected condition difference at baseline (Supplementary Table 5) despite the fact that participants were randomized by the study application and study staff were blinded to condition assignment (see the *Randomization* section in Supplementary Materials). To account for these baseline differences, we calculated change values between pre–mid and mid–post timespans. We then conducted a 2 (condition: slow-paced breathing, random-paced breathing) × 2 (timespan: mid–pre, post–mid) ANOVA with baseline Aβ values included as a covariate. For plasma Aβ42, there was a significant main effect of condition, *F*(1, 55) = 4.46, *p* = .039, *ηp²* = .075, indicating that the slow-paced breathing condition showed greater decreases (i.e., more negative mid–pre and post–mid change scores) than the random-paced breathing condition (Table 3). The covariate was also significant, *F(*1, 55) = 5.38, *p* = .024, *ηp²* = .089. There was no significant effect of timespan, *F*(1, 55) = 1.51, *p* = .225, *ηp²* = .025, nor a timespan × condition interaction, *F*(1, 55) = 0.02, *p* = .892, *ηp²* < .001, indicating that the condition differences in change were consistent across the two timespans.

**Table 3.** Mean change values and standard errors (in parentheses) of plasma Αβ levels.

| Plasma biomarker | Slow-paced breathing |  | Random-paced breathing |  | Effect of condition |
| --- | --- | --- | --- | --- | --- |
|  | mid-pre | post-mid | mid-pre | post-mid |  |
| Aβ42 | -0.393<br>(0.205) | -0.346<br>(0.295) | 0.053<br>(0.221) | 0.006<br>(0.318) | $F = 4.46$<br>$p = .039$<br>$\eta p^2 = .075$ |
| Aβ40 | -12.380<br>(7.727) | -0.045<br>(8.273) | -4.365<br>(8.310) | 1.723<br>(8.898) | $F = 1.01$<br>$p = .319$<br>$\eta p^2 = .018$ |
| Aβ42 & Aβ40<br>aggregate | -.056<br>(0.111) | -.039<br>(0.133) | 0.146<br>(0.119) | 0.011<br>(0.143) | $F = 2.65$<br>$p = .109$<br>$\eta p^2 = .046$ |

**Note:** Mean change values were adjusted for the covariates (i.e., baseline Aβ values).

For plasma Aβ40, there was no significant main effect of condition, *F*(1, 55) = 1.01, *p* = .319, *ηp²* = .018 (Table 3). The covariate was significant, *F*(1, 55) = 6.49, *p* = .014, *ηp²* = .105. There was no significant effect of timespan, *F*(1, 55) = 3.81, p = .056, *ηp²* = .065, nor a timespan × condition interaction, *F*(1, 55) = 0.08, *p* = .778, *ηp²* = .001.

There were no baseline differences in plasma Aβ42/40 ratios between conditions.

Therefore, we conducted a pre-registered 2 (condition) × 3 (time: pre, mid, post) ANOVA. There was no significant linear effects of time, *F*(1, 56) = 0.15, *p* = .701, *ηp²* = .003, condition, *F*(1, 56) = 1.51, *p* = .224, *ηp²* = .026, or time × condition interaction, *F*(1, 56) = 2.16, *p* = .147, *ηp²* = .037 (Supplementary Table 5).

### 3.3. PVS volume

For the CSO, there was no significant main effect of condition, *F*(1, 52) = 0.03, *p* = .874, *ηp²* < .001, or interaction between time and condition, *F*(2, 52) = 2.88, *p* = .096, *ηp²* = .052. There was a significant linear main effect of time, *F*(1, 52) = 4.33, *p* = .042, *ηp²* = .077. However, when we examined each condition separately, there was no significant linear effect of time in the slow-paced breathing condition, *F*(1, 28) = 0.35, *p* = .557, *ηp²* = .012, or in the random-paced breathing condition, *F*(1, 24) = 3.49, *p* = .074, *ηp²* = .127 (Supplementary Table 6). The results for other brain regions are also reported in Supplementary Table 6.

### 3.4. Hippocampal volume

To assess the test–retest reliability of hippocampal subfield volumes across the three time points (pre, mid, and post), intraclass correlation coefficients (ICCs) were computed using a two-way mixed-effects model with a consistency definition. As shown in *Supplementary Table 7*, ICC values (Average Measures) were consistently high across subfields in both the left and right hippocampus, with most values exceeding 0.95. These findings suggest a high level of measurement consistency in hippocampal subfield segmentation across time points.

We performed a two-way mixed ANCOVA (condition x time) on volume, including condition (slow-paced and random-paced) as a between-subject factor and time-point (pre, mid, and post) as a within-subject factor while controlling for intracranial volume as a covariate. For the left whole hippocampus, there was no significant interaction between time and condition, *F*(2, 104) = 1.08, *p* = .343, *ηp²* = .020, no main effect of time, *F*(2, 104) = 0.56, *p* = .573, *ηp²* = .011, or main effect of condition, *F*(1, 52) = 0.55, *p* = .464, *ηp²* = .010. For the right whole hippocampus, there was no significant interaction between time and condition, *F*(2, 104) = 0.38, *p* = .684, *ηp²* = .007, no main effect of time, *F*(2, 104) = 0.28, *p* = .758, *ηp²* = .005, or main effect of condition, *F*(1, 52) = 0.23, *p* = .637, *ηp²* = .004. Additionally, we applied the same two-way mixed ANCOVA model to the volumes of 19 hippocampal subfields in both the left and right hemispheres. None of the subfields showed a significant interaction effect between time and condition (all *p* >.072; Supplementary Table 8) .

### 3.5. Lumosity brain game training performance

Increments in game play performance over the course of the intervention were captured on latent level (first T-PLSC component, *p*permutation-corrected < 0.001; see Supplementary Figure 3a), with all games reliably contributing to the latent variable. Next, to evaluate longitudinal changes in cognitive performance during the intervention, a linear mixed-effects model was fitted to participants’ latent performance scores across the 10 time bins. The model included fixed effects for time, intervention group, their interaction, and baseline performance (time bin 1 latent score), with a random intercept for each participant.

There was a significant main effect of time (*β* = 0.46, 95% CI = [0.34, 0.57], *t*(595) = 7.94, *p* < .001), indicating a general increase in latent cognitive performance over the course of the 10-week period. Baseline performance was also a strong positive predictor of subsequent latent scores (*β* = 1.17, 95% CI = [1.10, 1.23], *t*(595) = 36.11, *p* < .001), as expected. However, there was no significant main effect of condition (*β* = −0.02, *t*(595) = −0.16, *p* = .87), and critically, no significant time × condition interaction (*β* < 0.001, *t*(595) = 0.004, *p* = .997). This suggests that the rate of cognitive improvement did not differ between the intervention groups after controlling for baseline performance. Similarly, time bin-wise comparisons did not reveal reliable group differences (all *p*s > 0.1; see Supplementary Figure 3b).

## 4. Discussion and conclusion

This ‘HeartBEAM’ clinical trial examined whether a 10-week slow-paced breathing intervention would influence plasma Aβ levels, PVS and hippocampal volumes, and cognitive performance (as assessed by Lumosity brain games), compared with a random-paced breathing control condition. The slow-paced breathing group showed greater decreases in plasma Aβ42 levels than the control group. No significant effects were observed for the other outcome measures. These non-significant findings may, in part, reflect methodological differences between the current study and our two previous clinical trials (i.e., the HRV-ER trial and the mindfulness trial; Min et al., 2023; Nashiro et al., 2025).

Specifically, there are two features that were shared across the other two clinical trials but differed in HeartBEAM. One is that in both earlier studies, the slow breathing was done at around a 10–13 seconds per breath pace. As described in the methods section, findings from our HRV-ER study indicated that breathing at rates of 11–13 seconds per breath was associated with greater reductions in plasma Aβ compared with faster rates (9–10 seconds). Consequently, in the current HeartBEAM study, we examined whether an even slower breathing rate of 15 seconds per breath could yield greater reductions in plasma Aβ levels. We determined for each participant in the slow breathing condition which pace elicited the largest heart rate oscillations, using multiple data points and statistical testing of confidence intervals rather than relying on numerically comparing single trials, as done in our previous study and in most HRV biofeedback studies (P. Lehrer et al., 2013). Based on assumptions in the HRV biofeedback field (P. Lehrer et al., 2013), we expected that we would find individual differences in which pace was best for each individual, influenced by their own estimated baroreflex frequency, or the low-frequency frequency at which they spontaneously showed the strongest heart-rate oscillatory dynamics even when not breathing at that frequency. However, the typical resting-state-estimated baroreflex frequency was around 0.1 Hz, but the 0.1 Hz breathing pace was not the most successful pace for increasing heart rate oscillations. We found that nearly everyone’s first best breathing pace was the slowest one from our initial set (15 seconds; Supplementary Figure 4). Across the practice weeks, as the application gave them new paces to try on either side of their best pace, participants’ best paces drifted even slower. Despite achieving large heart rate oscillations, the slow-paced breathing intervention yielded weaker Aβ effects than the slow breathing conditions in our previous HRV-ER and mindfulness trials. This suggests that simply maximizing heart rate oscillations is not the key factor in decreasing plasma Aβ; rather, breathing specifically at the baroreflex frequency may be critical for reducing plasma Aβ. The physiological mechanism may involve baroreflex stimulation, which simultaneously suppresses central noradrenergic activity and enhances cholinergic activity, both of which may reduce amyloidogenic processing of APP (Cavalcante et al., 2021; Heusser et al., 2010; Yao et al., 2022).

A second factor shared by the HRV-ER and the mindfulness trial is that in both, the interventions were framed as involving meditation. In HeartBEAM, the two conditions were introduced as involving paced breathing with no discussion of meditation. However, both HRV-ER and mindfulness clinical trials indicate simply attempting to meditate is not sufficient to reduce plasma Aβ. In both HRV-ER and the mindfulness study, the condition involving a meditative state without paced breathing increased plasma Aβ, while the condition involving a meditative state with paced breathing decreased plasma Aβ. Thus, our findings from the HRV-ER and mindfulness trials suggest that, for meditation to decrease plasma Aβ, it should be combined with slow-paced breathing.

It is important to address the study’s limitations. First, we observed unexpected baseline differences between conditions for both plasma Aβ42 and Aβ40 (Supplementary Table 5) despite participants being randomized by the study application and study staff being blinded to condition assignment. Second, this study didn’t account for lifestyle factors, such as sleep (Ju et al., 2017; Liu et al., 2020, 2022; Shokri-Kojori et al., 2018; Wei et al., 2017) and exercise (Feter et al., 2024; Rodriguez-Ayllon et al., 2024; Vasconcelos-Filho et al., 2021), which may influence Aβ levels. Future studies should include lifestyle factors known to influence Aβ levels as covariates. Third, future studies should incorporate additional physiological measures, particularly those that allow calculation of baroreflex sensitivity indices. This would help us better understand the role of the baroreflex in outcomes resulting from slow paced breathing.

In summary, the three completed clinical trials to date suggest that slow-paced breathing is most effective at reducing plasma Aβ when practiced in the context of a meditation session and at around 10–13 seconds per breath and that it was a limitation of the current study that participants were not attempting to meditate and that slow breathing was typically 15 seconds per breath or slower. Our upcoming clinical trial (NCT06410157; the Cardiac-Control Affecting Learning Through Mindfulness [CALM] project) addresses these potential limitations and examines whether baroreflex-frequency breathing during mindful attention to breath can influence PVS, hippocampal volume, and cognitive performance, in addition to plasma Aβ.

## Data Availability

All data produced in the present study are available upon publication.

## Acknowledgements

This research was funded by NIH R01AG080652 and Epstein Breakthrough Alzheimer’s Research Fund. During the work on this article, MJD was a member of the International Max Planck Research School on Computational Methods in Psychiatry and Ageing Research (IMPRS COMP2PSYCH, https://www.mps-uclcentre.mpg.de/comp2psych. Participating institutions: Max Planck Institute for Human Development, University College London). Acknowledgement is made to the donors of the ADR A2024006F, a program of the BrightFocus Foundation, for support of this research (MJD).

## Declaration of Generative AI and AI-assisted technologies in the writing process

ChatGPT was used solely to improve the readability of this manuscript. The authors reviewed and edited the content after using the tool and take full responsibility for the final version.

