## Supplementary Material for "Testing effects of paced breathing on plasma Aβ and brain perivascular spaces"

### Supplementary Materials

#### 1. Supplementary Methods

*1.1. Participant Compensation*

Participants received compensation for completing each week of home assessments, attending lab visits, completing the assigned intervention, and bonuses based on intervention performance. Participants earned $30 for each week of home assessments, $50 for each lab visit, $1 for each daily set of brain games, and $2 for completing both breathing practices daily. Bonuses could be earned based on performance on the brain games and breathing practices. Brain games bonuses were calculated weekly; if a participant's average score for the previous week was higher than the average score for plays prior to the previous week, they earned a $2 bonus. For breathing practice bonuses, participants received $1 when their average score from the daily 30-minute breathing practice was higher than the median of the scores from their previous three 15-minute breathing sessions.

*1.2. Randomization*

We created a blocked randomization scheme to assign participants to one of the two conditions: slow-paced breathing and random-paced breathing. When participants registered for our study application, they provided their sex (male or female). The study application counted the number of existing participants who were already assigned to a condition and who were the same sex as the incoming participant. If the number was even, the incoming participant was assigned to a randomly chosen condition. Otherwise, the study application found the condition of the most recently assigned participant of the same sex and assigned the incoming participant to the opposite condition. Researchers and participants were not informed of the assigned conditions to maintain blindness.

***Supplementary Figure 1:*** **Standardized Lumosity game performance over game plays**

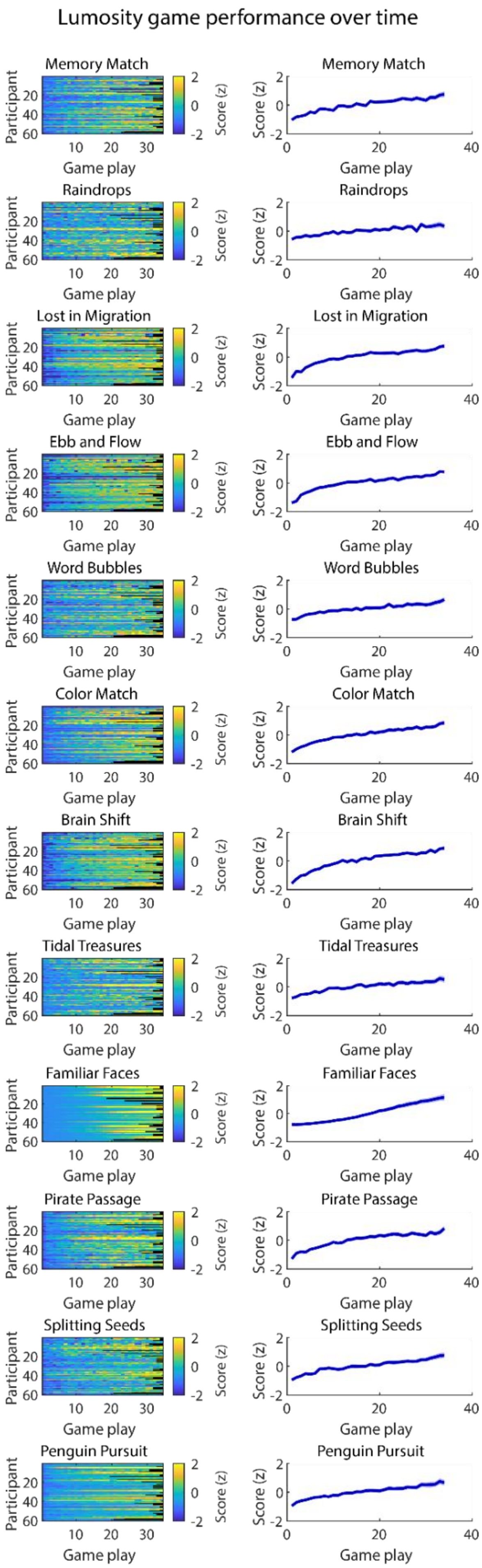
Lumosity game performance over time
Memory Match
Raindrops
Lost in Migration
Ebb and Flow
Word Bubbles
Color Match
Brain Shift
Tidal Treasures
Familiar Faces
Pirate Passage
Splitting Seeds
Penguin Pursuit
Participant
Game play
Score (z)

**Note:** Standardized game-wise performance time series for each participant (left) and averaged across participants (right). The Lumosity score performance metric was used for all analyses.

***Supplementary Figure 2:*** **Association of Lumosity performance across games**

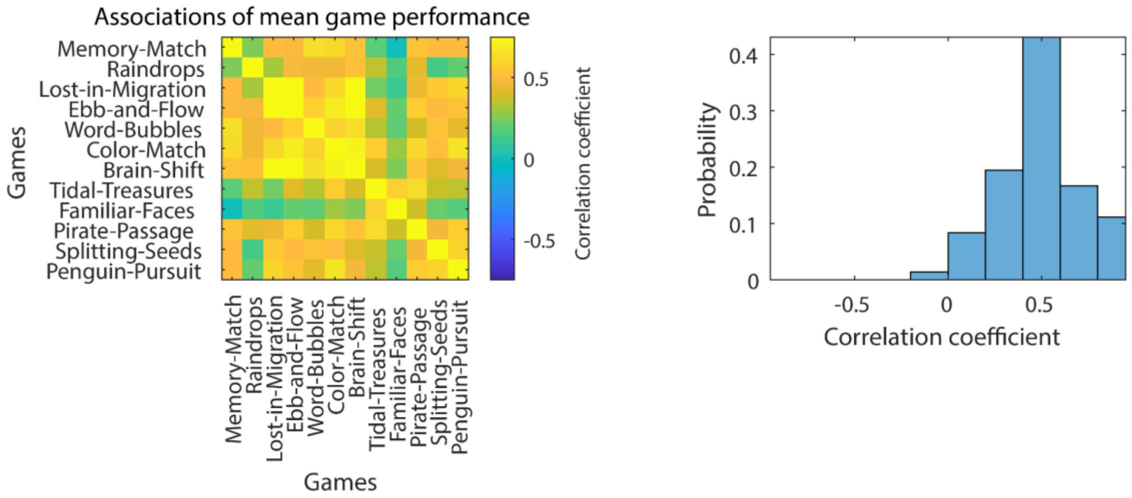


**Note:** Performance time series were averaged across time (within participants) before analysis.

***Supplementary Figure 3:*** Latent Lumosity game performance over time

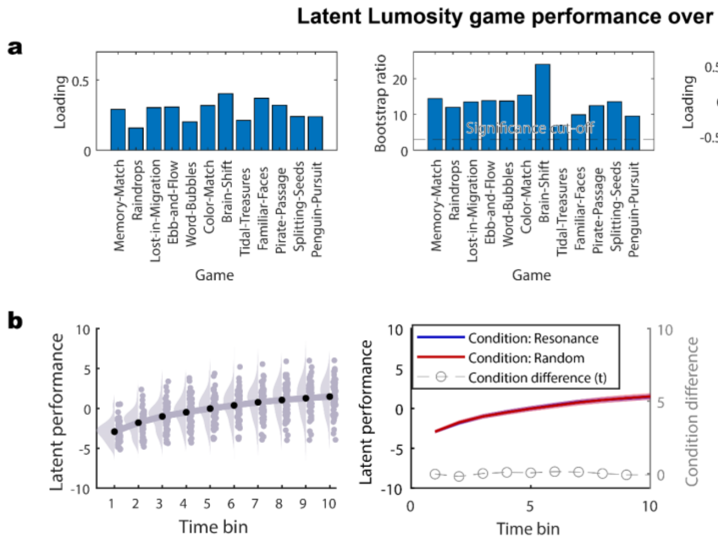

**Note:** Estimation of latent game performance trajectories using Task Partial Least Squares correlation (a) and condition differences in latent performance increments over the course of the study (b). Loadings/saliencies express how much a given task or time bin contribute to the latent association, while bootstrap ratios express the reliability of the contribution.

***Supplementary Figure 4:*** Average best breathing rates in the slow-paced breathing condition

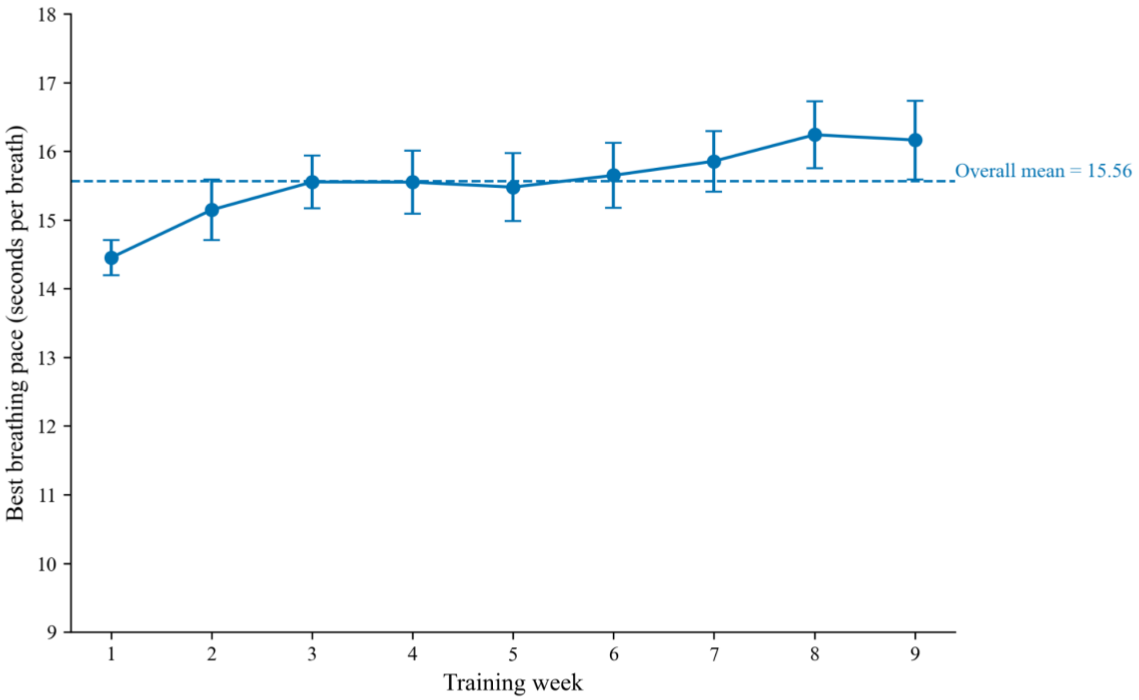


**Note:** In the slow-paced breathing condition in this study, nearly everyone's initial optimal rate was the slowest in our starting set (i.e., 15 seconds). Over time, as additional rates around their optimum were sampled, participants' best paces drifted even slower.

***Supplementary Table 1: Lumosity games***

| Set | Game name | Category | Description |
|---|---|---|---|

| | | | |
|---|---|---|---|
| 1 | Familiar faces | Memory | Participants play a waiter’s role and earn higher tips if they can remember their customers’ names and food orders |
| 1 | Tidal treasures | Memory | On each trial, participants are shown several unique ocean treasures and must choose one that they have not already selected in that round. Rounds can include up to 35 items, and some items are quite similar |
| 1 | Lost in migration | Attention | Participants indicate the direction of the central bird in the formation while ignoring the distractors around it |
| 1 | Splitting seeds | Attention | Participants evenly divide a pile of seeds without counting them |
| 1 | Pirate passage | Reasoning | Participants navigate their ship to reach the treasure island without colliding with other pirate ships |
| 1 | Ebb and flow | Flexibility | Participants view green or orange leaves moving across a pond and indicate the direction of where green leaves are pointing or where orange leaves are moving |
| 2 | Memory match | Memory | Participants must quickly determine whether a flashcard symbol matches the one presented two items ago |
| 2 | Word bubbles rising | Language | Participants generate as many words as possible that start with the same starting letters (e.g., res, medi) within the time limit |
| 2 | Raindrops | Math | Participants perform each math problem inside each raindrop before it reaches the bottom of the screen. Math problems include addition, subtraction, multiplication, and division |
| 2 | Penguin pursuit | Attention | Participants guide a penguin through a maze to reach a reward of fish at the end before the other penguin does. When the maze rotates, participants must rotate their mental map of the maze and recalibrate the directions to get to the goal |
| 2 | Brain shift | Flexibility | Participants are shown a letter-number pair (e.g., 5E) on the top or the bottom card. If the letter-number pair shows up on top, participants indicate whether the number is even; if it appears at the bottom, participants indicate whether the letter is a vowel or not |
| 2 | Color match 2 | Flexibility | Participants indicate the color of a written word while ignoring the meaning of the word |

**Note:** Game descriptions were retrieved from Nashiro et al. (2024).

Cognitive domains were retrieved from Steyvers and Schafer (2020).

***Supplementary Table 2: Regions comprising the centrum semiovale (CSO) using FreeSurfer parcellation mask***

| **Labels** | **ROI Name** |
|---|---|
| 3003 | wm-lh-caudalmiddlefrontal |
| 3008 | wm-lh-inferiorparietal |
| 3018 | wm-lh-parsopercularis |
| 3019 | wm-lh-parsorbitalis |
| 3020 | wm-lh-parstriangularis |
| 3022 | wm-lh-postcentral |
| 3024 | wm-lh-precentral |
| 3027 | wm-lh-rostralmiddlefrontal |
| 3028 | wm-lh-superiorfrontal |
| 3029 | wm-lh-superiorparietal |
| 3031 | wm-lh-supramarginal |
| 4003 | wm-rh-caudalmiddlefrontal |
| 4008 | wm-rh-inferiorparietal |
| 4018 | wm-rh-parsopercularis |
| 4019 | wm-rh-parsorbitalis |
| 4020 | wm-rh-parstriangularis |
| 4022 | wm-rh-postcentral |
| 4024 | wm-rh-precentral |
| 4027 | wm-rh-rostralmiddlefrontal |

| | |
|---|---|
| 4028 | wm-rh-superiorfrontal |
| 4029 | wm-rh-superiorparietal |
| 4031 | wm-rh-supramarginal |

**Note:** The white matter labels are based on Desikan-Killiany-Tourville adult cortical parcellation atlas (Klein & Tourville, 2012). The left hemisphere is denoted by 'wm-lh', and the right hemisphere is denoted by 'wm-rh'.

***Supplementary Table 3: Lumosity gameplays***

| Game | Memory Match | Raindrops | Lost in Migration | Ebb and Flow | Word Bubbles | Color Match | Brain Shift | Tidal Treasures | Familiar Faces | Pirate Passage | Splitting Seeds | Penguin Pursuit |
|---|---|---|---|---|---|---|---|---|---|---|---|---|
| min | 19 | 19 | 17 | 17 | 19 | 19 | 19 | 18 | 18 | 18 | 17 | 19 |
| max | 97 | 109 | 144 | 123 | 137 | 131 | 148 | 54 | 89 | 45 | 128 | 57 |
| mean | 33.6 | 34 | 34.7 | 34.3 | 34.6 | 34.6 | 34.6 | 33.1 | 33.9 | 32.8 | 34.2 | 32.8 |
| std | 9.2 | 10.7 | 15 | 12.4 | 14.3 | 13.5 | 15.5 | 4.8 | 8.4 | 4.3 | 13 | 5.1 |
| median | 34 | 34 | 34 | 34 | 34 | 34 | 34 | 34 | 34 | 34 | 34 | 34 |

***Supplementary Table 4: Means and standard errors (in parentheses) of HRV indices by condition and state (baseline resting, training)***

| HRV index | Slow-paced breathing | | Within-group state effect | Random-paced breathing | | Within-group state effect | Condition | State | State x Condition |
|---|---|---|---|---|---|---|---|---|---|
| | rest | training | *p* | rest | training | *p* | *p* | *p* | *p* |
| Heart rate | 74.83(1.65) | 76.89(1.65) | 0.058 | 74.10(1.68) | 74.94(1.68) | 0.727 | 0.561 | 0.013 | 0.290 |
| Log RMSSD | 3.78(0.07) | 4.05(0.07) | <0.001 | 3.89(0.08) | 4.17(0.08) | <0.001 | 0.260 | <0.001 | 0.902 |
| Log HF power | 5.34(0.14) | 5.91(0.14) | <0.001 | 5.45(0.14) | 6.26(0.14) | <0.001 | 0.205 | <0.001 | 0.153 |
| Log LF power | 6.03(0.16) | 7.56(0.16) | <0.001 | 6.01(0.16) | 6.85(0.16) | <0.001 | 0.067 | <0.001 | 0.007 |
| Log coherence | 0.82(0.04) | 1.37(0.04) | <0.001 | 0.71(0.04) | 0.67(0.04) | 0.857 | <0.001 | <0.001 | <0.001 |

**Note:** Values are presented as means with standard errors (SE) in parentheses. Normality was assessed using the Shapiro–Wilk test. Except for heart rate, all HRV indices violated normality assumptions and were therefore natural log–transformed prior to analysis. Within-group state effects (rest vs. training) were tested using paired t tests. Effects of condition, state, and their interaction were tested using linear mixed-effects models (LMMs). $p < .05$ was considered statistically significant. Heart rate is presented in beats per minute (bpm).

***Supplementary Table 5: Means and standard errors (in parentheses) of plasma Aβ levels by condition and time point***

| Plasma biomarker | Slow-paced breathing | | | Random-paced breathing | | | Effect of condition | Linear effect of time | Time x condition interaction |
|---|---|---|---|---|---|---|---|---|---|
| time | pre | mid | post | pre | mid | post | | | |
| Aβ42 | 10.263<br>(0.370) | 10.037<br>(0.356) | 9.692<br>(0.377) | 12.106<br>(0.396) | 11.967<br>(0.381) | 11.971<br>(0.404) | $F = 16.43$<br>$p < .001$<br>$\eta p^2 = .227$ | $F = 3.91$<br>$p = .053$<br>$\eta p^2 = .065$ | $F = 1.49$<br>$p = .227$<br>$\eta p^2 = .026$ |
| Aβ40 | 272.246<br>(8.950) | 265.226<br>(9.476) | 263.507<br>(9.229) | 305.390<br>(9.590) | 294.870<br>(10.154) | 298.516<br>(9.889) | $F = 7.52$<br>$p = .008$<br>$\eta p^2 = .118$ | $F = 2.62$<br>$p = .111$<br>$\eta p^2 = .045$ | $F = 0.04$<br>$p = .847$<br>$\eta p^2 = .001$ |
| Aβ42 & Aβ40 aggregate | -0.371<br>(0.150) | -0.332<br>(0.151) | -0.387<br>(0.150) | 0.343<br>(0.161) | 0.381<br>(0.161) | 0.410<br>(0.161) | $F = 14.04$<br>$p < .001$<br>$\eta p^2 = .200$ | $F = 0.11$<br>$p = .737$<br>$\eta p^2 = .002$ | $F = 0.31$<br>$p = .580$<br>$\eta p^2 = .006$ |
| Aβ42/40 ratio | 0.038<br>(0.001) | 0.039<br>(0.001) | 0.037<br>(0.001) | 0.040<br>(0.001) | 0.041<br>(0.001) | 0.040<br>(0.001) | $F = 1.51$<br>$p = .224$<br>$\eta p^2 = .026$ | $F = 0.15$<br>$p = .701$<br>$\eta p^2 = .003$ | $F = 2.16$<br>$p = .147$<br>$\eta p^2 = .037$ |

**Note:** Aβ aggregate scores were calculated as the average of the Z-scores of Aβ42 and Aβ40

***Supplementary Table 6: Means of PVS volume fractions in 8 brain regions for each condition and time point***

| Brain region | Slow-paced breathing | | | Within-group time effect | Random-paced breathing | | | Within-group time effect | Condition | Time | Time x Condition |
|---|---|---|---|---|---|---|---|---|---|---|---|
| | pre | mid | post | *p* | pre | mid | post | *p* | *p* | *p* | *p* |
| CSO | 0.0079(.0043) | 0.0088(.0052) | 0.0080(.0039) | 0.557 | 0.0077(.0034) | 0.0083(.0042) | 0.0092(.0054) | 0.074 | 0.874 | 0.042* | 0.096 |
| middle temporal | 0.0052(.0035) | 0.0053(.0031) | 0.0049(.0030) | 0.287 | 0.0052(.0029) | 0.0057(.0033) | 0.0061(.0038) | 0.093 | 0.530 | 0.244 | 0.037* |
| entorhinal | 0.0021(.0017) | 0.0028(.0030) | 0.0032(.0030) | 0.042* | 0.0027(.0024) | 0.0025(.0023) | 0.0029(.0038) | 0.809 | 0.953 | 0.151 | 0.298 |
| parahippocampal | 0.0041(.0027) | 0.0040(.0030) | 0.0036(.0023) | 0.266 | 0.0038(.0029) | 0.0032(.0022) | 0.0033(.0022) | 0.311 | 0.467 | 0.132 | 0.990 |
| superior temporal | 0.0041(.0032) | 0.0047(.0039) | 0.0043(.0029) | 0.509 | 0.0046(.0027) | 0.0047(.0030) | 0.0052(.0038) | 0.334 | 0.536 | 0.225 | 0.495 |
| medial orbitofrontal | 0.0063(.0039) | 0.0083(.0083) | 0.0061(.0043) | 0.821 | 0.0058(.0038) | 0.0070(.0071) | 0.0082(.0094) | 0.135 | 0.933 | 0.190 | 0.129 |
| fusiform | 0.0037(.0028) | 0.0043(.0036) | 0.0038(.0027) | 0.451 | 0.0045(.0026) | 0.0043(.0023) | 0.0043(.0025) | 0.699 | 0.524 | 0.894 | 0.518 |
| basal ganglia | 0.00068(.00027) | 0.00066(.00025) | 0.00064(.00027) | 0.327 | 0.00065(.00026) | 0.00068(.00031) | 0.00071(.00026) | 0.218 | 0.779 | 0.849 | 0.117 |

**Note:** There was no significant interaction between time and condition in any region except for middle temporal regions. For the middle temporal regions, neither the main effect of condition,

$F(1, 52) = 0.40$, $p = .530$, $\eta p^2 = .008$, nor the linear effect of time, $F(1, 52) = 1.39$, $p = .244$, $\eta p^2 = .026$, was significant. However, there was a significant interaction between time and condition, $F(1, 52) = 4.58$, $p = .037$, $\eta p^2 = .081$. Standard deviations are shown in parentheses.

***Supplementary Table** 7*

*Intraclass Correlation Coefficients (ICCs) for Test–Retest Reliability of Hippocampal Subfield Volumes Across Three Time Points*

| | Hemisphere | | | | | | | | | |
|---|---|---|---|---|---|---|---|---|---|---|
| | Left | | | | | Right | | | | |
| region | ICC (Average Measures) | 95% CI Lower | 95% CI Upper | F (54,108) | p | ICC (Average Measures) | 95% CI Lower | 95% CI Upper | F (54,108) | p |
| Hippocampal tail | 0.993 | 0.99 | 0.996 | 150.32 | < .001 | 0.988 | 0.98 | 0.992 | 80.19 | < .001 |
| subiculum-body | 0.983 | 0.973 | 0.989 | 57.98 | < .001 | 0.984 | 0.974 | 0.99 | 61.32 | < .001 |
| CA1-body | 0.985 | 0.977 | 0.991 | 68.48 | < .001 | 0.983 | 0.973 | 0.989 | 58.42 | < .001 |
| subiculum-head | 0.976 | 0.963 | 0.985 | 41.87 | < .001 | 0.98 | 0.969 | 0.988 | 50.37 | < .001 |
| hippocampal-fissure | 0.92 | 0.875 | 0.951 | 12.47 | < .001 | 0.96 | 0.937 | 0.975 | 24.80 | < .001 |
| presubiculum-head | 0.939 | 0.905 | 0.963 | 16.49 | < .001 | 0.899 | 0.841 | 0.937 | 9.86 | < .001 |
| CA1-head | 0.984 | 0.975 | 0.99 | 61.48 | < .001 | 0.988 | 0.982 | 0.993 | 85.43 | < .001 |
| presubiculum-body | 0.976 | 0.963 | 0.985 | 41.73 | < .001 | 0.972 | 0.957 | 0.983 | 36.02 | < .001 |
| parasubiculum | 0.983 | 0.974 | 0.99 | 59.36 | < .001 | 0.985 | 0.977 | 0.991 | 68.29 | < .001 |
| molecular layer HP-head | 0.924 | 0.881 | 0.953 | 13.11 | < .001 | 0.93 | 0.89 | 0.957 | 14.28 | < .001 |
| molecular layer HP-body | 0.911 | 0.861 | 0.945 | 11.24 | < .001 | 0.944 | 0.912 | 0.966 | 17.86 | < .001 |
| GC-ML-DG-head | 0.984 | 0.974 | 0.99 | 61.15 | < .001 | 0.986 | 0.978 | 0.991 | 69.80 | < .001 |
| CA3-body | 0.968 | 0.951 | 0.981 | 31.72 | < .001 | 0.965 | 0.945 | 0.978 | 28.29 | < .001 |
| GC-ML-DG-body | 0.98 | 0.968 | 0.987 | 49.13 | < .001 | 0.975 | 0.961 | 0.985 | 40.48 | < .001 |
| CA4-head | 0.982 | 0.972 | 0.989 | 56.83 | < .001 | 0.986 | 0.978 | 0.991 | 70.03 | < .001 |
| CA4-body | 0.979 | 0.967 | 0.987 | 47.10 | < .001 | 0.979 | 0.967 | 0.987 | 47.25 | < .001 |
| fimbria | 0.958 | 0.934 | 0.974 | 23.62 | < .001 | 0.968 | 0.95 | 0.98 | 31.40 | < .001 |
| CA3-head | 0.983 | 0.974 | 0.99 | 59.24 | < .001 | 0.977 | 0.964 | 0.986 | 43.33 | < .001 |
| HATA | 0.983 | 0.973 | 0.989 | 57.30 | < .001 | 0.988 | 0.98 | 0.992 | 80.14 | < .001 |
| Whole hippocampal body | 0.988 | 0.981 | 0.993 | 83.82 | < .001 | 0.991 | 0.985 | 0.994 | 106.13 | < .001 |
| Whole hippocampal head | 0.993 | 0.989 | 0.996 | 139.37 | < .001 | 0.994 | 0.991 | 0.996 | 170.52 | < .001 |
| Whole hippocampus | 0.994 | 0.991 | 0.996 | 173.79 | < .001 | 0.996 | 0.993 | 0.997 | 232.60 | < .001 |

***Supplementary Table 8***

*Results of 2-Way ANCOVAs on Hippocampal Subfield Volumes Showing Time × Condition Interaction Effects with ICV as a Covariate*

| | Hemisphere | | | | | |
|---|---|---|---|---|---|---|
| | Left | | | Right | | |
| region | *F* | *p* | *ηp²* | *F* | *p* | *ηp²* |
| Hippocampal tail | 0.684 | .507 | .013 | .465 | .629 | .009 |
| subiculum-body | 2.694 | .072 | .049 | 1.616 | .204 | .030 |
| CA1-body | 1.142 | .323 | .021 | .381 | .684 | .007 |
| subiculum-head | .049 | .952 | .001 | .098 | .907 | .002 |
| hippocampal-fissure | .558 | .515 | .013 | 1.059 | .350 | .020 |
| presubiculum-head | .536 | .587 | .010 | .466 | .629 | .009 |
| CA1-head | 1.093 | .339 | .021 | .417 | .660 | .008 |
| presubiculum-body | .890 | .414 | .017 | .934 | .396 | .018 |
| parasubiculum | .755 | .472 | .014 | .213 | .809 | .004 |
| molecular layer HP-head | .090 | .914 | .002 | .318 | .728 | .006 |
| molecular layer HP-body | .794 | .455 | .015 | 1.0 | .371 | .019 |
| GC-ML-DG-head | .541 | .584 | .010 | .009 | .991 | .0 |
| CA3-body | .177 | .838 | .003 | .729 | .485 | .014 |
| GC-ML-DG-body | .830 | .439 | .016 | .049 | .952 | .001 |
| CA4-head | .098 | .906 | .002 | .003 | .997 | .0 |
| CA4-body | .218 | .804 | .004 | .302 | .740 | .006 |
| fimbria | 2.476 | .089 | .045 | .482 | .619 | .009 |
| CA3-head | .346 | .709 | .007 | .031 | .969 | .001 |
| HATA | .070 | .932 | .001 | .620 | .540 | .012 |
| Whole hippocampal body | 1.544 | .218 | .029 | .722 | .488 | .014 |
| Whole hippocampal head | .634 | .533 | .012 | .338 | .714 | .006 |
| Whole hippocampus | 1.082 | .343 | .020 | .382 | .684 | .007 |

**Note:** Each ANCOVA tested the interaction between time (pre, mid, post) and condition ( slow-paced, random-paced), controlling for intracranial volume (ICV). Only interaction effects are reported (F, p, partial $\eta^2$).
